# A prevalence study of Autism Spectrum Disorder in the Russian Federation

**DOI:** 10.1101/2025.10.20.25338431

**Authors:** Oksana I. Talantseva, Raisa S. Romanova, Julia E. Kuznetsova, Viktoria A. Manasevich, Katerina V. Lind, Mariia A. Parfenenko, Julia Benoit, Elena L. Grigorenko

## Abstract

Autism Spectrum Disorder (ASD) is a neurodevelopmental condition marked by heterogeneity in presentation. In recent decades, ASD prevalence has risen globally, yet data from non-Western and middle-income countries, including Russia, remain scarce. Official statistics report a prevalence of 0.41 per 1000 (0.041%), far below global estimates of 32.2 per 1000 (3.2%), indicating underdiagnosis. This study provides the first population-based estimate of ASD prevalence among Russian elementary school children and examines barriers to diagnosis and treatment. The study followed a two-phase epidemiological approach. ASD identification used standardized instruments (SCQ, ADOS-2, ADI-R). The target population included all students in grades 1–3 (n = 34 847), stratified into mainstream, special education, and resource classes. ASD prevalence was estimated using Bayesian regression correcting for screening misclassification. The estimated ASD prevalence was 22.2 per 1000 children (95% CIs: 18.6–36.0), substantially exceeding what is reported based on the RF administrative data. The diagnostic phase had a 25.6% participation rate. ASD was more frequently identified in students from special education and resource classes than in general education settings. A Bayesian sensitivity analysis confirmed the robustness of prevalence estimates across plausible values for sensitivity and specificity. These findings suggest that ASD prevalence in Russia is considerably underestimated via administrative data. Systemic barriers—including limited access to diagnostic services and stigma surrounding psychiatric labels—may hinder ASD identification. Addressing these challenges is essential for improving early detection and service provision in Russia and similar contexts.

## Introduction

Autism Spectrum Disorder (ASD) is a lifelong neurodevelopmental condition characterized by social communication impairments and restricted and/or repetitive behaviors and interests [1,2]. Given the astronomical diversity of manifestations in these domains, divergences are marked by a striking heterogeneity in ASD presentation [3]. This heterogeneity is further compounded by wide variability in intellectual and language functioning, intra-individual cognitive discrepancies [4,5], and frequent comorbidity with other developmental and psychiatric conditions [6]. Together, these factors place ASD among the most disabling developmental disorders, imposing a substantial economic burden and necessitating diverse health service utilization [7].

Since the 1970s, ASD prevalence studies have been conducted in at least 37 countries— primarily high-income ones—while data from many low- and middle-income countries remain scarce [8]. These studies have demonstrated a substantial increase in ASD prevalence estimates worldwide since the late 20th century. Notably, data from the Autism and Developmental Disabilities Monitoring (ADDM) Network of the USA CDC highlight this trend, with prevalence rates among American 8-year-olds rising from 6.7 per 1,000 in 2000 [9] to 32.2 per 1,000 in 2022 [10], indicating a 380.6 % increase.

Although the reasons for the reported upward trend remain under discussion, a substantial body of research highlights that reported prevalence estimates do not necessarily reflect true rates. Instead, these estimates are shaped by multiple factors, including the progression and refinement of diagnostic criteria; advancements in screening; diagnostic and policy practices; increased awareness and advocacy for ASD; potential environmental alterations, and the quality of research [11]. Furthermore, prevalence estimates vary by case ascertainment methods and geographic regions, adding to the complexity of accurately measuring true ASD rates. Specifically, a recent meta-analysis [12] identified geographical region and socio-economic status as significant moderators. The highest estimates were reported for high-income countries, while the lowest - for low-income ones. Even within the same high-income country and within a single study encompassing different regions, these discrepancies persist. For instance, the most recent ADDM Network estimates ranged from 9.7/1,000 in Texas to 53.1/1,000 in California, despite the network’s ongoing and increasingly successful efforts to reduce regional discrepancies through improved policies [10].

In Russia, psychiatric diagnoses are still based on ICD-10, under which ASD is considered an umbrella term for the following formal diagnostic codes: F84.0, F84.1, F84.4, F84.5, F84.8, F84.9. To date, no targeted research has been conducted on the prevalence of ASD in RF, with the sole exception of a study that assessed ASD risk using a domestic screening tool [13] in a sample of children aged 18 to 48 months across nine regions. This study estimated the risk of ASD at 1.8 per 1,000 (0.18%), which is substantially lower than risk estimates obtained using established screening instruments worldwide [14]. The latest available statistics of the State Psychiatric Service in 2022 [15] have indicated a prevalence of ASD in Russia at 0·41 cases per 1,000, or at 0.041% (0.40 cases of childhood and atypical autism and 0.007 cases of Asperger’s syndrome). Among individuals officially diagnosed with autism, 78.2% were reported to have a disability, with 94.7% of these being children under 17. Compared with 2018 (0.32 cases per 1,000, or 0.03%), the estimated prevalence of ASD has increased by 85.5%, likely reflecting improvements in diagnostic approaches and detection of ASD within the country. A more prominent increase from 2018 to 2020 was registered among children (aged 0–14 years) and adolescents (aged 15–17 years): from 0.11 to 0.17 cases and from 0.06 to 0.13 cases per 1,000, respectively. A retrospective study analyzing trends in governmental statistics on registered cases of ASD has also revealed a steady upward trend — critically delayed compared to worldwide estimates — observed in Russia since 2014 [16]. Thus, despite positive trends, the prevalence of ASD remains critically low — approximately 29 times lower than the conservative global median rate [11]. Thus, it is likely that a significant proportion of people with autism in Russia do not receive a diagnosis and, consequently, cannot access services. Consistent with other nationally representative samples, significant regional differences have also been observed, though these appear substantially uneven in their distribution. In 2021, estimates ranged from 0.17/10,000 in the Kaluga region to 17.77/10,000 cases in Yamalo-Nenets Autonomous Okrug [16]. Given this disparity, autistic individuals and their families may find themselves in a challenging situation, often having to relocate in pursuit of an accurate diagnosis and appropriate services.

Here we report the first population-based prevalence study in Russia. We targeted the entire elementary school population in a large city, covering both a mainstream population sample and groups with a high risk of ASD. We also provide available administrative data reporting officially diagnosed ASD cases in the school-based population.

## Materials and Methods

### Study Site

The study was conducted in a large city that serves as the administrative center of one of Russia’s 89 federal subjects, located in the Central Federal District (one of eight federal districts in the country). As of January 1, 2023, this federal subject (oblast) had a population of 2,285,300, with the city itself accounting for 1,061,108 residents. To date, this city is considered one of Russia’s more progressive cities in terms of state-supported ASD-related services. Since 2013, the interdepartmental project *Autism: Routes of Assistance* has established early ASD risk screening, diagnostic services, and structured educational programs in the city. Hence, we selected this site as it ensures access to state-funded ASD care. In other words, children who received an ASD diagnosis in this study were referred to local providers for clinical confirmation and services.

### Study Population

The target population consisted of all 1^st^-, 2^nd^-, and 3^rd^-grade students attending mainstream or special education schools in the city during the 2023–2024 academic year (N = 34,847; 17,732 boys). The target population was divided into three separate strata, depending on the risk of having ASD. Different sampling strategies were employed for each stratum:

#### (1) Low-Risk (LR) Subpopulation

The LR target subpopulation (N = 34,392) comprised all children attending mainstream elementary schools (N = 118), except for schools that participated in the pilot study (N = 11). Given the large number of students in this subpopulation and the fact that a number (N = 18) of mainstream schools have resource classes for children with ASD, we used a mixed sampling strategy. First, all schools with resource classes were invited to participate in the study. Then, we randomly selected schools without resource classes based on the assumption that together, they would provide an adequate representation of all schools. The final sample size was determined based on a judgment of the optimal balance between the minimum sufficient sample size estimate, the research team’s limited resources, and results of the pilot study. Overall, 30 schools were selected (N = 8,565; 51.7% boys), covering 24.9% of the LR target subpopulation.

#### (2) Elevated-Risk (ER) Subpopulation

The ER subpopulation included all elementary students attending special education schools (N = 5), implementing programs for children with developmental disabilities (N = 292; 65.4% boys). Given the significant challenges in ASD diagnostics in Russia and the tendency of psychiatrists to assign only a single diagnosis without considering potential comorbidities [16], we assumed that this subpopulation might have an increased risk of ASD as well as possibly a misdiagnosis. Due to the relatively small size of this subpopulation, all relevant schools were invited to participate in the study. Thus, a convenience sampling design was applied.

#### (3) High-Risk (HR) Subpopulation

All children enrolled in resource classes (N = 112; 70.5% boys) were classified within the HR stratum. Since enrollment in resource classes requires both a medical and an educational diagnosis of ASD, we considered the risk of ASD in this subpopulation to be extremely high—approaching 100%. A convenience sampling design was also applied within the HR population: all schools were sought as eligible and invited to participate, and all children and their parents were recruited.

A detailed breakdown of the total number of schools in the study cite, including their distribution by type and participation status, is provided in Supplementary Table 1.

### Case Identification

To identify cases of ASD, we applied a two-phase design—screening and identification of children at risk for ASD, followed by clinical assessment and diagnostic confirmation of those who screened positive—regardless of strata (Figure 1).

**Figure 1.**
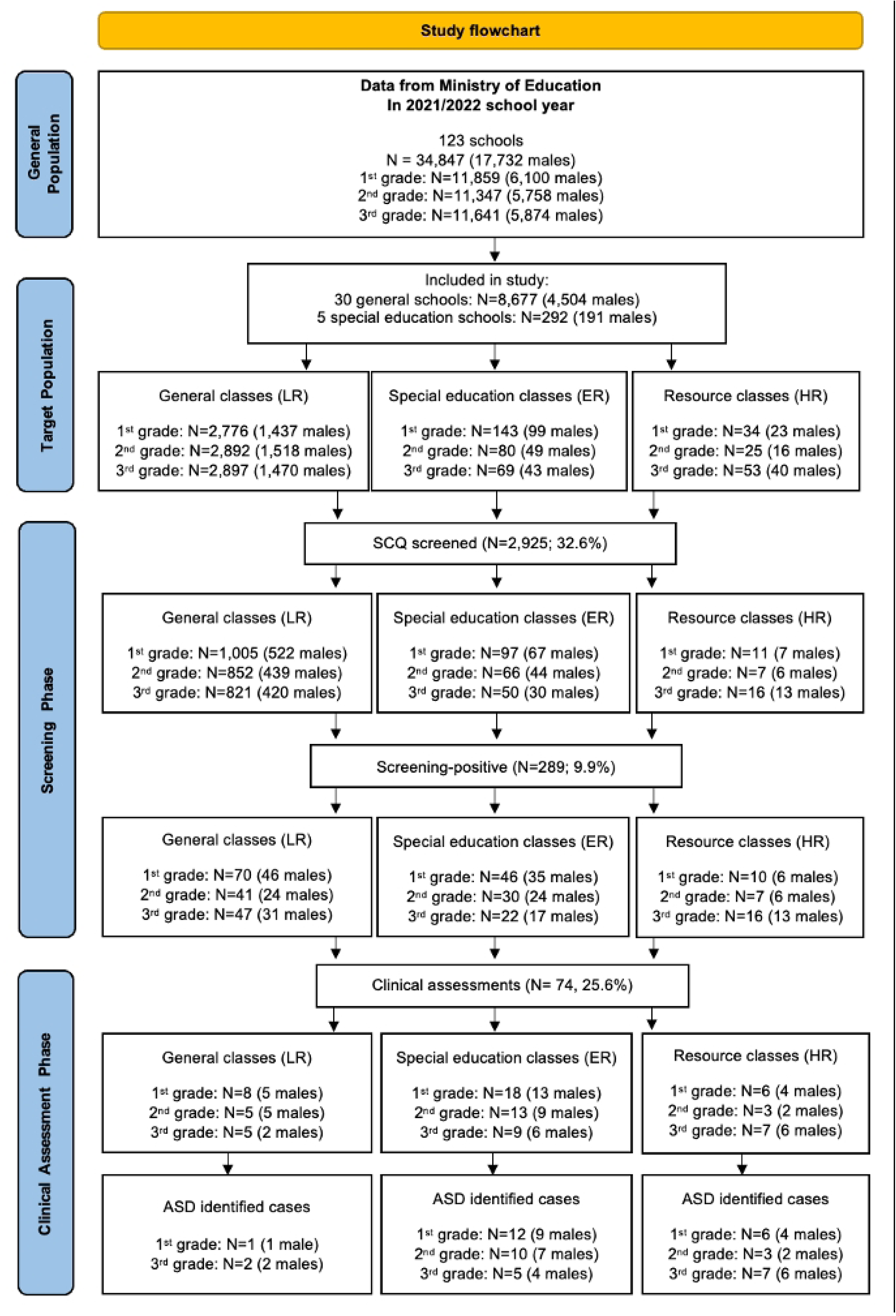
Flow-chart describing the case identification process across the 2 phases of the study.

#### (1) Screening Phase

This phase was carried out from January 2023 to May 2024. To increase participation, pediatricians serving the selected schools were invited to act as interviewers during the first phase. Their on-site work was managed by a designated research team member, whose responsibilities included communicating with school principals, obtaining permission for school participation, and organizing the storage and delivery of completed packages of documents to the research team. Parent meetings were held separately at each school. Pediatricians attended these meetings, explained the aims of the study, distributed screening packages, encouraged participation, and answered any questions that arose. All participating pediatricians were trained by the research team and received monetary compensation for their support of the study.

Each envelope contained two identical informed consent forms (one copy was for the parents to keep), a consent form for personal data processing, and a screening questionnaire (Social-Communication Questionnaire, SCQ, Lifetime version) [17]. Each form had an ID sticker; the SCQ forms did not include any identification information. Parents who declined to participate in the screening or did not sign the informed consent were considered non-respondents.

All these forms were completed at administrative meetings. The completed envelopes were returned to the research team for data entry, which was carried out with integrated quality control procedures. Randomly selected ten percent of the data were double-entered, and another ten percent of SCQ forms were checked for consistency against the printed copies.

Participants who scored ≥11 on the SCQ were categorized as screening-positives. Although a cutoff score of 15 is typically recommended to define the at-risk group, we used a threshold of 11. This decision was based on previous studies suggesting that a score between 11 and 13 is more optimal for epidemiological research, as it reduces the number of false negatives [18]. This choice was also influenced by the fact that the Russian-language version of the SCQ has not yet been validated on a sufficiently large sample, indicating that its optimal cutoff score may differ from that of the original version. Initially, we planned to sample both screening-positive and screening-negative groups with unequal probabilities during the Diagnostic Confirmation Phase, in order to estimate the proportion of ASD cases in each group and incorporate these estimates into the prevalence analysis, adjusted for non-response. However, constraints related to study duration, funding, and organizational challenges prevented us from following this protocol. As a result, only screening-positive cases were eligible for recruitment in the second phase.

#### (2) Diagnostic Confirmation Phase

This phase was carried out between July 2023 and November 2024. All children who screened positive and met eligibility criteria (availability of informed consent and contact information), along with their parents, were invited to participate in a multidisciplinary diagnostic confirmation evaluation. The screening consent form for personal data processing included a field for providing a phone number to contact the child’s parent. Extensive efforts were made to reach the parents of all eligible children. To issue the invitation, the research team made several contact attempts, including two text messages sent several days apart, each containing an introduction and an invitation to participate. If there was no response, this was followed by two calls. Additional exclusion criteria at this step included: (1) no phone number; (2) an incorrect or disconnected number; and (3) no response to phone calls or text messages after four contact attempts by the recruiter.

Evaluation of the autism-related features was performed using The Autism Diagnostic Observation Schedule, Second Edition (ADOS-2) [19] and The Autism Diagnostic Interview - Revised (ADI-R) [20]. Cognitive development was assessed via The Leiter International Performance Scale, Third Edition (Leiter-3) [21], adaptive functioning – through The Vineland Adaptive Behavior Scales, Second Edition (Vineland-II) [22]. Multiple visits were required to complete this battery. During the first visit, the research team provided parents with detailed information about all planned procedures, and parents were asked to sign an informed consent to participate in the second phase of the study.

According to the original study protocol, a comprehensive diagnostic battery was planned to be used. However, in the pilot study, the number of refusals to participate increased with each additional assessment method. As a result, the ADOS-2 and SCQ-retest were made mandatory, while the remaining assessments were made voluntary.

All clinical assessments were conducted by the research team and administered at government and private centers that provide services for children with ASD. Upon parental request, the research team also provided written feedback, including descriptions of the child’s test scores and behaviors.

The final diagnostic decision was made in two steps. In the first step, the results of the assessment instruments were coded separately, and a preliminary diagnosis was made based on whether the child met the algorithm criteria of the diagnostic assessments. A diagnosis of ASD was given if the threshold score was met or exceeded, and symptoms were present across all relevant diagnostic domains. In the second step, the case was reviewed by considering the results of both assessments together. If there was a discrepancy between ADOS-2 and ADI-R outcomes, a consensus diagnosis was reached in conjunction with the clinical judgment of the research team, following DSM-5-TR diagnostic criteria.

Detailed information on all screening and diagnostic instruments used during both phases of the study is provided in the Supplemental Appendix.

### Statistical Analysis

To assess potential bias due to nonparticipation in the second phase, we first examined predictors of participation among screen-positive children. We hypothesized that families differ in their underlying willingness to pursue diagnostic evaluation, and that this latent trait may be partially explained by observable characteristics. Using logistic regression, we found that SCQ scores were significantly associated with participation (*p* < .05), whereas sex and age were not (*p* = .83 and *p* = .76, respectively). Higher SCQ scores—particularly those above the conventional cutoff—were associated with increased likelihood of attendance. This finding justified the inclusion of the SCQ score as a covariate in subsequent models.

To estimate ASD prevalence while accounting for missing diagnostic data and misclassification, we implemented a Bayesian logistic regression model in which ASD status was treated as a latent binary variable. Observed diagnostic outcomes were modeled as error-prone measurements of this latent status, conditional on known values of screening sensitivity (Se) and specificity (Sp). The probability of a positive diagnosis was thus a function of both the unobserved true ASD status and the psychometric properties of the screening tool, following established frameworks for probabilistic bias analysis [23] and Bayesian modelling of misclassified outcomes [24]. The individual probability of true ASD was modeled as a function of SCQ score.

In the primary analysis, Se and Sp were fixed at 0.85 and 0.75, respectively, based on published validation studies.^19^ The model was implemented in R using the *rjags* package [25] with three MCMC chains, 5,000 burn-in, and 10,000 post-burn-in iterations. Posterior ASD probabilities were aggregated within each stratum (LR, ER, HR) and combined using known population sizes to produce post-stratified prevalence estimates. Posterior distributions were summarized using median and 95% Bayesian credible intervals (CrIs).

To assess robustness, we conducted deterministic sensitivity analysis across 16 plausible combinations of Se (0.80–0.95) and Sp (0.65–0.80), based on values reported in previous SCQ validation studies [16,17]. Additionally, a probabilistic sensitivity analysis treated Se and Sp as uncertain parameters with informative Beta priors (Se ∼ Beta(85,15); Sp ∼ Beta(60,40)). We also explored the influence of assumptions in the HR stratum by constraining its posterior prevalence to 100%, 95%, 90%, or 85%. Full model details are provided in the Supplementary Appendix.

### Administrative Prevalence

In addition to the main study, we also requested statistical data on diagnoses compatible with ASD (ICD-10 codes F84.0, F84.1, F84.4, F84.5, F84.8, and F84.9) that were registered in the city’s educational system among elementary school students from 2020 to 2024. Prevalence estimates were calculated as simple proportions, dividing the number of cases by the size of the target population.

## Ethical Approval

The study was conducted in accordance with the Declaration of Helsinki and was approved by the Bioethics Committee of Sirius University of Science and Technology (date of approval: April 15, 2021). The study protocol was submitted to the Department of Health of the participating city in April of 2021, which provided organizational support and coordination for the research through a network of the city’s pediatric clinics. Additionally, the study involved active participation from the federal subject’s Ministry of Education, the city’s Department of Education, and the principals of both regular and specialized schools.

## Results

Thirty-five schools participated (including five special education schools and ten with resource classes), yielding a total target population of 8,976 children. The overall response rate was 32.6% in the screening phase and 25·6% in the diagnostic phase, with lower participation in mainstream settings. Participation challenges were linked to the COVID-19 pandemic and the proximity of the study region to the front lines of the Russian-Ukrainian conflict. The flow of participants through the study is described in Figure 1.

The pooled prevalence adjusted for participation bias was 15.9 per 1,000. Bayesian correction for misclassification yielded a post-stratified prevalence of 22.2 per 1,000 (95% CrI: 18.6–36.0), with adjusted sex-specific rates of 24.7 per 1,000 in males and 17.7 per 1,000 in females. The empirical sex ratio of 3.2:1 was reduced to 1.4:1 after adjustment. Detailed prevalence results are provided in Table 1.

**Table 1.** ASD Prevalence Estimates by Sex and Grade.

|  | Population |  | N cases |  | Sex Ratio | Prognosed cases |  | Prevalence (95% CrIs) per 1,000 |  | Overall |
| --- | --- | --- | --- | --- | --- | --- | --- | --- | --- | --- |
|  | M | F | M | F |  | M | F | M | F |  |
| 1<sup>st</sup> Grade | 596 | 517 | 14 | 5 | 2.8 | 14 | 9 | 25.6 (20.5; 45.4) | 17.5 (15.5; 30.6) | 22.4 (18.6; 38.5) |
| 2<sup>nd</sup> Grade | 489 | 436 | 9 | 4 | 2.3 | 10 | 10 | 22.5 (18.9; 32.7) | 24.3 (19.9; 33.5) | 23.3 (19.9; 32.9) |
| 3<sup>rd</sup> Grade | 462 | 425 | 12 | 2 | 6.0 | 12 | 4 | 26.0 (21.6; 45.2) | 13.9 (9.7; 28.0) | 20.7 (16.7; 36.2) |
| Overall | 1547 | 1378 | 35 | 11 | 3.2 | 36 | 22 | 24.7 (20.3; 40.8) | 17.7 (14.5; 29.5) | 22.2 (18.6; 36.0) |

ASD = autism spectrum disorder; M = males; F = females; CrIs = Credible Intervals

Sensitivity analyses confirmed robustness of prevalence estimates across a range of plausible screening sensitivity and specificity values, as well as under conservative assumptions in the HR stratum (see Figure 2 and 3, and Supplementary Appendix for full details).

**Fig. 2.**
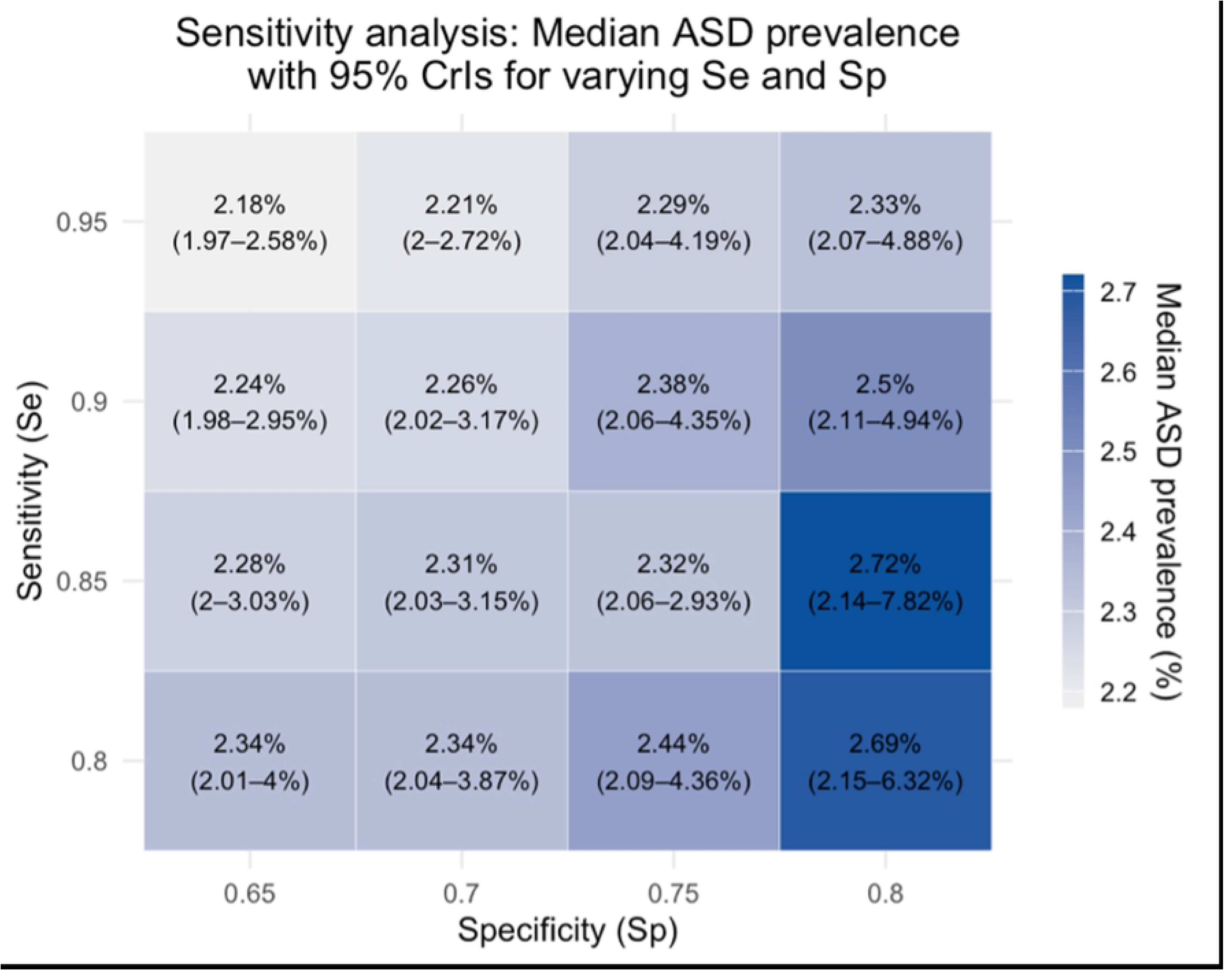
Post-stratified ASD prevalence estimates under varying screening sensitivity (Se) and specificity (Sp). Each cell shows the median ASD prevalence (%) obtained from deterministic sensitivity analysis using a Bayesian logistic regression model with corresponding 95% credible intervals shown in parentheses.

**Fig. 3.**
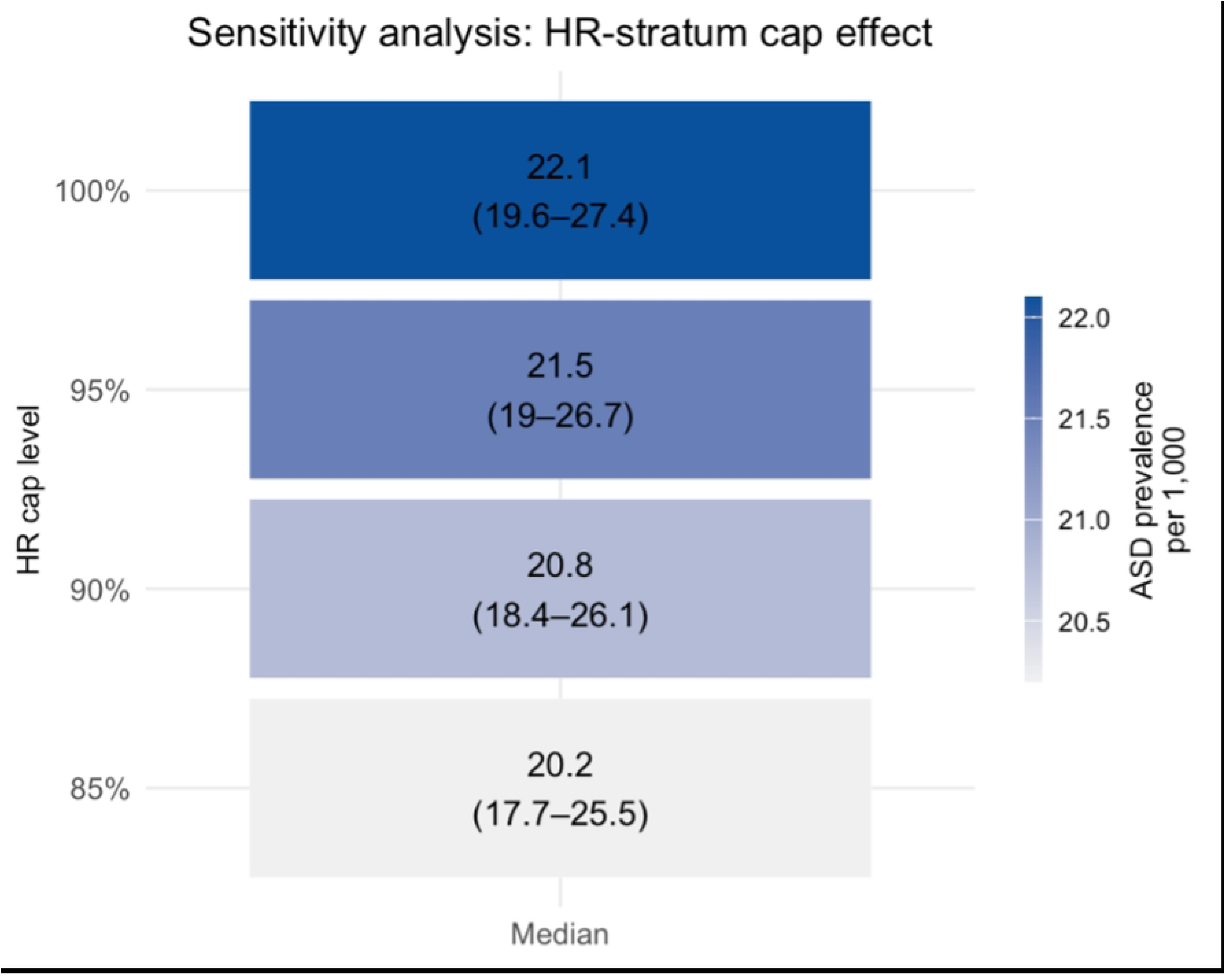
Sensitivity analysis of post-stratified ASD prevalence under varying upper bounds for the HR stratum. Each row represents a model scenario in which the predicted prevalence in the high-risk (HR) stratum was artificially capped at a maximum value (100%, 95%, 90%, or 85%). Cells show the median ASD prevalence per 1000, post-stratified across HR, IR, and LR strata. Color intensity reflects the magnitude of prevalence. The results show minimal variation (20.4–20.6 per 1000), supporting the robustness of the overall estimate.

Among participants diagnosed with ASD, intellectual functioning was assessed in 20 children. In the LR stratum, the children demonstrated average IQ (M = 101.5, SD = 0.7). In the ER stratum, 30% had average or above-average IQ, 30% were in the borderline range, and 40% met criteria for mild intellectual disability (M = 76.4, SD = 16.5). In the HR strata, one child (12.5%) had an average IQ, 50% were in the borderline range, and 37.5% had mild intellectual disability (M = 75.6, SD = 9.9).

Adaptive functioning was evaluated in 24 participants using the Vineland-II. In the LR group, one child had age-appropriate adaptive behavior, while the other showed mild deficits. In the ER group, one child (7.7%) had adequate functioning, two (15.4%) were borderline, and the majority (76.9%) demonstrated impairments, including one case with moderate deficits. In the HR group, one participant showed borderline functioning, and the remaining eight of nine children (88.9%) had impaired adaptive behavior, ranging from mild to moderate severity.

The raw prevalence of ASD, which is calculated based on data obtained from official educational sources for the 2023-2024 academic year, was 4.1 cases per 1,000. Similar estimates (Table 2) were also observed for the two preceding academic years (4.1/1000 and 4.0/1000 in 2021-2022 and 2022-2023, respectively). In 2020-2021, the prevalence was slightly higher—5.0 cases per 1,000.

**Table 2.**
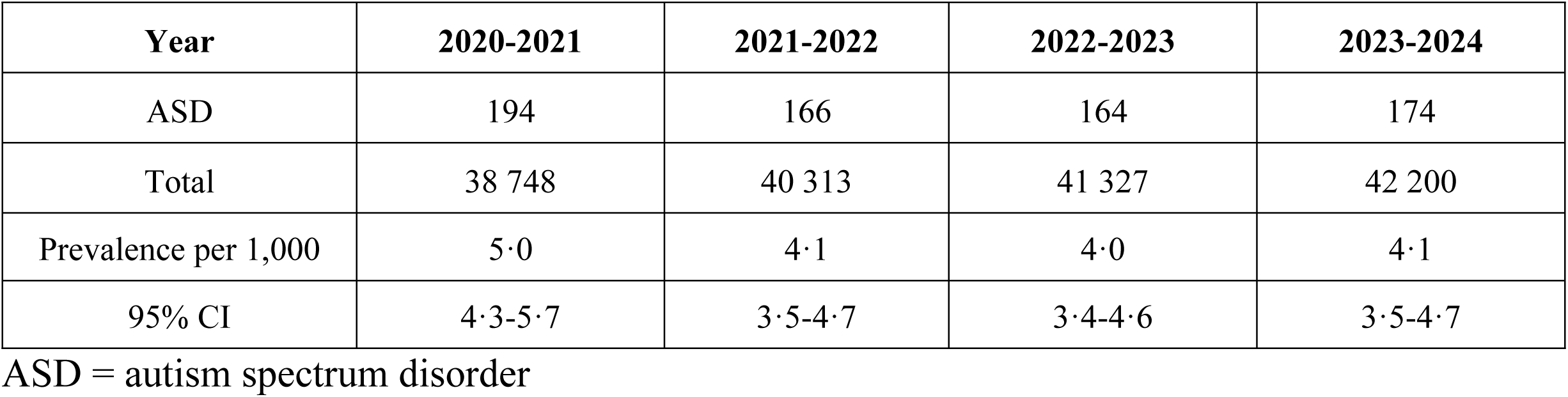
Administrative ASD Prevalence.

## Discussion

To our knowledge, this is the first population-based prevalence study of ASD in Russia. Based on direct clinical assessment, the estimated prevalence of ASD among elementary school students was 2.2%. This estimate exceeds the global consensus rate of 1% accepted by the World Health Organization [26] and is higher than summary estimates reported in recent meta-analyses [11, 12]. At the same time, our findings are consistent with more recent international studies, many of which report prevalence rates approaching or exceeding 2%. For example, a recently published study reported an ASD prevalence in Cyprus at 1.8% [27]. The result is comparable with the estimate reported in a study with a similar analysis about the prevalence of ASD in South Korea [28]. In comparison, our estimates fall below those from the U.S.-based ADDM Network, which reported prevalence rates of 2.8% and 3.2% in 2020 and 2022, respectively [11, 29]. This discrepancy may reflect differences in methodology, as ADDM studies have been shown to yield higher estimates relative to population-based designs that rely on direct clinical evaluation [12].

The empirically observed male-to-female ratio in ASD prevalence was 3.2:1, aligning with recent literature that challenges the long-standing ratio of 4.0–4.5:1. Our findings support the revised estimate of approximately 3.0:1, as reported by Loomes et al. [30] and other recent studies accounting for diagnostic biases that disproportionately affect girls [11, 31]. Notably, when adjusting for multiple potential biases using Bayesian hierarchical modeling—including screening misclassification, non-response, and sample structure—the male-to-female ratio in prevalence was further reduced to 1.4:1.

The prevalence derived from administrative data in the Russian educational system was substantially lower, ranging from 0.4% to 0.5% between 2020 and 2024. On the one hand, this discrepancy provides strong evidence of underdiagnosis in the school-age population and highlights the need to improve diagnostic services and awareness of ASD in Russia. On the other hand, such discrepancies between administrative and population-based estimates are common across countries [32], especially when administrative data rely on passive detection or incomplete records.

The current study has several limitations. The most notable is the low response rate, which may have introduced bias into the results. Despite substantial organizational support from the local Ministries of Health and Education, as well as from school principals and teachers, many parents were reluctant or unwilling to participate. While in the first phase, we collected approximately 50% of the distributed SCQ questionnaires, participation in the second phase did not exceed 30%. According to the research team and regional partners, a significant barrier to family participation was the study’s focus on identifying ASD as a psychiatric diagnosis. Stigmatization of individuals with mental health conditions, reluctance to be associated with the issue (e.g., ‘I have a healthy child,’ ‘This doesn’t concern us’), especially when it involves a child’s health—and a general lack of experience with research participation all influenced decisions regarding consent. Indeed, parents of children attending specialized schools—who were already aware of their children’s health conditions—were more likely to respond to the research team’s invitations, particularly when there was uncertainty about the formal diagnosis. In contrast, families of children in resource classes were often less motivated to participate due to a well-established diagnosis.

Despite these limitations, we believe that our results, interpreted with appropriate caution, offer important insights into the burden of ASD in Russia. They can inform health system planning and resource allocation, highlight regional disparities in diagnostic coverage, and guide future epidemiological efforts to identify social or environmental risk factors. As the first study of its kind in the Russian context, these findings help fill a geographic gap in global ASD prevalence research and contribute essential data to future meta-analyses and global surveillance efforts.

## Data Availability

The datasets used and analyzed during the current study available from the corresponding author on reasonable request. All R scripts used for Bayesian prevalence estimation, including model specification, post-stratification, and sensitivity analyses (deterministic and probabilistic), are available at: https://github.com/OksanaTalantseva/prevalence-asd-russia/tree/main

## Acknowledgments

We express our sincere gratitude for their assistance in organizing the study to the Ministry of Health and the Ministry of Education of the Voronezh Region, the Department of Education and Youth Policy of the Administration of the Urban District of Voronezh, We would also like to thank Chief Pediatrician Margarita Kinshina and Director of the Centre for Psychological, Pedagogical, Medical and Social Assistance to Families Olga Nedorezova, pediatricians and nurses at children’s clinics, teachers and school principals in Voronezh.

We wish to extend our heartfelt gratitude to the Autism-Regions Association, a large-scale association comprising 92 parent organizations in 33 regions of Russia. We are indebted to the Association’s activity and efforts, which were instrumental in initiating the first Russian study of the prevalence of ASD in children. Special thanks go to the children and parents who participated in the study and made it possible.

We are grateful to our scientific advisor, Eric Fombonne, psychiatrist, epidemiologist, head of autism research at the Institute for Development and Disability at Oregon Health & Science University, for his invaluable contributions to the execution and analysis of the study.

Also, we are grateful to Lauren Elderton from the University of Houston for her editorial assistance.

The views expressed in this article are those of the authors and do not necessarily reflect the opinions of the organizations, professionals, and families who assisted in the organization and conduct of the research.

## Contributors

OIT – contributed to developing the concept, study design, and protocols, conducted and supervised clinical evaluations, conducted statistical analysis of the prevalence estimates, drafted/wrote and reviewed the manuscript. RSR – contributed to the developing study protocols, recruited and evaluated the patients, supervised clinical evaluations, managed data, conducted descriptive statistical analysis, drafted/wrote and reviewed the manuscript. JEK – managed the research project, drafted/wrote the manuscript. VAM - recruited and evaluated the patients, supervised clinical evaluations, conducted descriptive statistical analysis, drafted/wrote the manuscript. EVL - contributed to the data analysis, reviewed the manuscript. JB – supervised the developing study design and data analysis, reviewed and edited the manuscript. MAP - reviewed the manuscript. ELG – developed the concept and study design, supervised clinical evaluations, reviewed and edited the manuscript.

